# Local patterns of genetic sharing challenge the boundaries between neuropsychiatric and insulin resistance-related conditions

**DOI:** 10.1101/2024.03.07.24303921

**Authors:** Giuseppe Fanelli, Barbara Franke, Chiara Fabbri, Josefin Werme, Izel Erdogan, Ward De Witte, Geert Poelmans, I. Hyun Ruisch, Lianne Maria Reus, Veerle van Gils, Willemijn J. Jansen, Stephanie J.B. Vos, Kazi Asraful Alam, Aurora Martinez, Jan Haavik, Theresa Wimberley, Søren Dalsgaard, Ábel Fóthi, Csaba Barta, Fernando Fernandez-Aranda, Susana Jimenez-Murcia, Simone Berkel, Silke Matura, Jordi Salas-Salvadó, Martina Arenella, Alessandro Serretti, Nina Roth Mota, Janita Bralten

## Abstract

The co-occurrence of insulin resistance (IR)-related metabolic conditions with neuropsychiatric disorders is a complex public health challenge. Evidence of the genetic links between these phenotypes is emerging, but little is currently known about the genomic regions and biological functions that are involved. To address this, we performed Local Analysis of [co]Variant Association (LAVA) using large-scale (N=9,725-933,970) genome-wide association studies (GWASs) results for three IR-related conditions (type 2 diabetes mellitus, obesity, and metabolic syndrome) and nine neuropsychiatric disorders. Subsequently, positional and expression quantitative trait locus (eQTL)-based gene mapping and downstream functional genomic analyses were performed on the significant loci. Patterns of negative and positive local genetic correlations (|r_g_|=0.21-1, p_FDR_<0.05) were identified at 109 unique genomic regions across all phenotype pairs. Local correlations emerged even in the absence of global genetic correlations between IR-related conditions and Alzheimer’s disease, bipolar disorder, and Tourette’s syndrome. Genes mapped to the correlated regions showed enrichment in biological pathways integral to immune-inflammatory function, vesicle trafficking, insulin signalling, oxygen transport, and lipid metabolism. Colocalisation analyses further prioritised 10 genetically correlated regions for likely harbouring shared causal variants, displaying high deleterious or regulatory potential. These variants were found within or in close proximity to genes, such as *SLC39A8* and *HLA-DRB1*, that can be targeted by supplements and already known drugs, including omega-3/6 fatty acids, immunomodulatory, antihypertensive, and cholesterol-lowering drugs. Overall, our findings underscore the complex genetic landscape of IR-neuropsychiatric multimorbidity, advocating for an integrated disease model and offering novel insights for research and treatment strategies in this domain.

**Highlights:**

- Local genetic correlations found even in the absence of global correlations.
- Both positive and negative local correlations found for IR-neuropsychiatric pairs.
- Enrichment for immune, and insulin signalling pathways, among others.
- Pinpointed shared likely causal variants within 10 genomic regions.
- Identified therapeutic targets, e.g., SLC39A8 and HLA-DRB1, for drug repurposing.

## 1. Introduction

Multimorbidity, defined as the co-occurrence of multiple conditions within an individual, poses substantial challenges to healthcare systems (Skou et al., 2022). An example is the observed co-occurrence of insulin resistance (IR)-related metabolic conditions, such as type 2 diabetes mellitus (T2DM), obesity, and metabolic syndrome (MetS), with neuropsychiatric disorders (Wimberley et al., 2022). This multimorbidity contributes to more severe physical and mental health outcomes, leading to reduced treatment effectiveness and higher mortality rates (Fanelli and Serretti, 2022; Kraus et al., 2023; Possidente et al., 2023). Moreover, IR is associated with detrimental effects on cognitive function, potentially worsening the cognitive impairment observed in various neuropsychiatric disorders (Fanelli et al., 2022b).

IR manifests as reduced tissue responsiveness to insulin stimulation, primarily disrupting blood glucose homeostasis and inducing long-term complications in the microcirculation and peripheral nervous system (DeFronzo et al., 2015). Such a metabolic perturbation is a distinctive feature of T2DM, central obesity, and MetS (DeFronzo et al., 2015). Emerging evidence suggests that IR shares aetiological pathways with neuropsychiatric disorders, including Alzheimer’s disease (AD), mood and psychotic disorders (Fanelli et al., 2022a; Hubel et al., 2019; Watson et al., 2019). The connection between IR-related conditions and neuropsychiatric disorders is supported by compelling epidemiological data (Leutner et al., 2023; Wimberley et al., 2022). Indeed, bidirectional phenotypic associations have been found between these two nosological groups (Wimberley et al., 2022). This evidence blurs the boundaries between traditional disease categories, advocating for a more integrated approach to research and clinical management (Chwastiak et al., 2015; Fanelli and Serretti, 2022). Consequently, a deeper comprehension of the mechanisms underlying this multimorbidity is essential.

Beyond shared environmental risk factors – including poor diet, sedentary lifestyle, and disturbed sleep (Marx et al., 2017; Ogilvie and Patel, 2018; Schuch et al., 2018), which could also be direct manifestations of psychopathology – shared genetic components have been identified (Fanelli et al., 2022a). Both IR-related conditions and neuropsychiatric disorders are highly heritable and polygenic (Mahajan et al., 2022; Trubetskoy et al., 2022), with heritability estimates, derived from twin and family studies, ranging from 40 to 80% (Almgren et al., 2011; Wray et al., 2014). Work by us and others disclosed global genetic correlations between neuropsychiatric disorders and IR-related conditions, indicative of shared genetic bases (Fanelli et al., 2022a; Hubel et al., 2019), though the effect directions were not consistent across all phenotype pairs. Intriguingly, two clusters of neuropsychiatric disorders were identified, wherein the genetics of IR-related conditions showed opposite directions of genetic correlation. The first included attention-deficit/hyperactivity disorder (ADHD) and major depressive disorder (MDD), which showed positive genetic correlations with IR-related conditions; the second included obsessive-compulsive disorder (OCD), anorexia nervosa (AN), and schizophrenia, which showed negative genetic correlations with IR-related conditions (Fanelli et al., 2022a). Genetic covariance was also highlighted within gene sets pertinent to insulin processing, secretion, and signalling, suggesting that several neuropsychiatric disorders could be reconceptualised as “insulinopathies” of the brain (Fanelli et al., 2022a). Strikingly, certain neuropsychiatric disorders, such as AD and bipolar disorder (BD), demonstrated no global genetic correlations with IR-related conditions, despite previous literature suggested a shared pathophysiology (Fanelli et al., 2022a; Shieh et al., 2020). However, global genetic correlation only encapsulates the average direction of genetic sharing across the genome, while the patterns of genetic correlations at the level of individual genomic regions can vary significantly (van Rheenen et al., 2019). Local genetic correlation can deviate from the genome-wide average, and regions of strong, local genetic correlation have been reported for multiple traits even in the absence of genome-wide correlation (van Rheenen et al., 2019; Werme et al., 2022). Therefore, the absence of genome-wide genetic correlations does not necessarily exclude shared genetics in specific regions, suggesting the importance to further study the possible genetic overlap between conditions without global genetic correlation, such as AD and IR-related traits (Fanelli et al., 2022a). Importantly, dissecting the local patterns of genetic sharing could shed light on specific genetic factors involved in IR-neuropsychiatric multimorbidity and new potential therapeutic targets for both groups of conditions. Recent bioinformatics advances have enabled more detailed exploration of the genetic overlap across phenotypes. Traditional global genetic correlation methods, like Linkage Disequilibrium Score regression (LDSC), assess shared genetic architecture between phenotypes across the entire genome (Bulik-Sullivan et al., 2015) but may fail in identifying phenotype pairs of that share individual genomic regions (Bulik-Sullivan et al., 2015). Therefore, the utilisation of local genetic correlation analyses may offer more granular insights into shared genetic bases (Werme et al., 2022).

In this study, we aimed to dissect the genetic overlap between three IR-related metabolic conditions – namely, obesity, T2DM, and MetS - and nine psychiatric disorders by examining their pairwise patterns of local genetic correlation throughout semi-independent regions across the genome. Any shared genomic region was further explored using positional and expression quantitative trait locus (eQTL)-based gene mapping techniques. This was followed by a functional annotation of the mapped genes, enabling a deeper exploration of biological mechanisms underlying IR-neuropsychiatric multimorbidity. Lastly, we investigated the shared (likely) causal variants possibly driving the pathophysiology of this comorbidity.

## 2. Material and methods

### 2.1. Input datasets

We leveraged publicly available summary statistics from the largest genome-wide association studies (GWASs) on the three most prevalent IR-related conditions, namely obesity, MetS, and T2DM (n=244,890-933,970), and nine neuropsychiatric disorders, including AD, ADHD, AN, autism spectrum disorder (ASD), BD, MDD, OCD, schizophrenia, and Tourette’s syndrome (TS) (n=9,725-933,970). These neuropsychiatric disorders were chosen because they are the best genetically characterised by the Psychiatric Genomics Consortium (Cross-Disorder Group of the Psychiatric Genomics Consortium. Electronic address and Cross-Disorder Group of the Psychiatric Genomics, 2019). Further details, including sample size of each GWAS, are reported in **Table 1**. To maintain consistency in genetic data, analyses were confined to individuals of European ancestry, employing the human genome build GRCh37/hg19 as a reference. All statistical analyses were performed using R v4.2.1 (2022-06-23).

**Table 1.** Characteristics of genome-wide association study (GWAS) samples used as input for Local Analysis of [Co]variant Association (LAVA) and follow-up genomic analyses included in this study. Abbreviations: MetS metabolic syndrome, T2DM type 2 diabetes mellitus, AD Alzheimer’s disease, ADHD attention-deficit/ hyperactivity disorder, AN anorexia nervosa, ASD autism spectrum disorder, BD bipolar disorder, MDD major depressive disorder, OCD obsessive-compulsive disorder, IOCDF-GC/OCGAS International OCD Foundation Genetics Collaborative/OCD Collaborative Genetics Association Studies, TS Tourette’s syndrome, PMID PubMed ID, N total sample size, Neff effective sample size [Neff = 4/(1/Cases + 1/Controls)].

| Phenotype | Authors | Year | PMID | Ancestry | N | Cases | Controls | N<sub>eff</sub> |
| --- | --- | --- | --- | --- | --- | --- | --- | --- |
| **MetS** | Lind | 2019 | 31589552 | European | 291,107 | 59,677 | 231,430 | 189,772.81 |
| **Obesity** | Watanabe et al. | 2019 | 31427789 | European | 244,890 | 9,805 | 235,085 | 37,649.69 |
| **T2DM** | Mahajan et al. | 2022 | 35551307 | European | 933,970 | 80,154 | 853,816 | 293,100.50 |
| **AD** | Wightman et al. | 2021 | 34493870 | European | 762,917 | 86,531 | 676,386 | 306,866.18 |
| **ADHD** | Demontis et al. | 2023 | 36702997 | European | 225,534 | 38,691 | 186,843 | 128,213.80 |
| **AN** | Watson et al. | 2019 | 31308545 | European | 72,517 | 16,992 | 55,525 | 52,041.91 |
| **ASD** | Grove et al. | 2019 | 30804558 | European | 46,350 | 18,381 | 27,969 | 44,366.62 |
| **BD** | Mullins et al. | 2021 | 34002096 | European | 413,466 | 41,917 | 371,549 | 150,669.89 |
| **OCD** | IOCDF-GC/OCGAS | 2018 | 28761083 | European | 9,725 | 2,688 | 7,037 | 7,780.14 |
| **MDD** | Howard et al. | 2019 | 30718901 | European | 500,199 | 170,756 | 329,443 | 449,855.91 |
| **Schizophrenia** | Trubetskoy et al. | 2022 | 35396580 | European | 130,644 | 53,386 | 77,258 | 126,281.98 |
| **TS** | Yu et al. | 2019 | 30818990 | European | 14,307 | 4,819 | 9,488 | 12,783.30 |

### 2.2. Local genetic correlation analyses

We utilised the R package LAVA (Local Analysis of [co]Variant Association) (https://github.com/josefin-werme/LAVA) to perform pairwise local genetic correlation analyses between the three IR-related conditions and the nine neuropsychiatric disorders (Werme et al., 2022). Compared to traditional global correlation analysis methods (Bulik-Sullivan et al., 2015), LAVA estimates the genetic correlation at smaller genomic loci, which provides a more fine-grained overview of the genetic overlap between traits. In addition to providing insight into the potentially heterogeneous nature of the shared association patterns across the genome, LAVA allows identification of the regions from which the pleiotropy is originating (Werme et al., 2022). Further details regarding the LAVA analytical steps are provided in the **Supplementary information** (**paragraph 1.1**). Given the total number of bivariate tests performed across all phenotype pairs, local genetic correlations were deemed as statistically significant at a maximum acceptable false discovery rate (FDR) of q=0.05, following the approach of Hindley et al. (2022).

### 2.3. Positional and eQTL gene mapping

The biomaRt R package (version 2.54.1) (https://doi.org/doi:10.18129/B9.bioc.biomaRt) (Durinck et al., 2005) was used to annotate single-nucleotide polymorphisms (SNPs) within each genetically correlated region and positionally map them to genes. We used the Ensembl database (release 109, GRCh37/hg19, *homo sapiens*) as a reference for gene annotations. We defined filters to specify the genomic regions of interest based on their location (chromosome number, start and end positions).

For the eQTL-based gene mapping, the loci2path R package (version 1.3.1) (https://doi.org/10.18129/B9.bioc.loci2path) (Xu et al., 2020) was used to identify eQTLs within the genetically correlated regions that may influence gene expression in 13 cortical, subcortical, and cerebellar brain regions (i.e., total brain cortex, frontal cortex BA9, hippocampus, hypothalamus, amygdala, anterior cingulate cortex BA24, caudate, nucleus accumbens, putamen, cervical spinal cord, substantia nigra, cerebellar hemisphere, cerebellum). We obtained the eQTL data from the Genotype-Tissue Expression (GTEx) project (GTEx V8, GRCh38/hg38) (https://gtexportal.org/home/dataset) and restricted our analysis to brain tissues due to their relevance to neuropsychiatric disorders. Prior to the analysis, we lifted the eQTL coordinates to the GRCh37/hg19 genomic build using the UCSC LiftOver tool (https://genome-store.ucsc.edu) to align with the used GWAS summary statistics.

### 2.4. Functional annotation of genetically correlated regions

Functional annotation analyses were conducted separately for each phenotype pair where genetically correlated regions were found. We employed the GENE2FUNC module within the Functional Mapping and Annotation of Genome-Wide Association Studies (FUMA) platform (Watanabe et al., 2017), using default parameters and multiple testing correction (Watanabe et al., 2017). This approach served to examine important properties of the mapped genes, such as their tissue-specific and temporal expression profiles, enrichment in predefined gene sets, potential as drug targets, and previous trait/disease associations. Detailed information on the methods applied for these analyses are presented in the **Supplementary information** (**paragraph 1.2**).

To contextualise our findings within the broader landscape of known disease associations, we also investigated the overrepresentation of the identified genes within those previously associated with traits or diseases by querying the NHGRI-EBI GWAS Catalog (Buniello et al., 2019).

### 2.5. Colocalisation analyses

To identify the specific shared causal variants within each region showing local genetic correlation, we conducted robust Bayesian colocalisation analyses through the coloc R package (Giambartolomei et al., 2014) and the Sum of Single Effects (SuSiE) regression framework (Wallace, 2021) (https://chr1swallace.github.io/coloc/articles/a06_SuSiE.html). Notably, these approaches allow for simultaneous evaluation of multiple causal genetic variants within a genomic region and are therefore not limited by the single causal variant assumption that traditional colocalisation methods use. The input genomic regions were those showing evidence of local genetic correlation between each pair of IR-related condition and neuropsychiatric disorder. The detailed methodology is reported in the **Supplementary information** (**paragraph 1.3**).

### 2.6. Functional annotation of 95% credible sets of shared causal variants

We employed the SNPnexus web server (https://www.snp-nexus.org/) to further characterise the functional significance of the likely causal variants identified by colocalisation (Oscanoa et al., 2020). This tool integrates a wealth of genomic and functional annotation resources to elucidate the potential biological consequences of variants on gene structure, regulation, and function. The analysis encompassed several annotation categories, including gene annotations, regulatory elements (e.g., miRBASE, CpG islands), and non-coding scoring (i.e., deleteriousness Combined Annotation Dependent Depletion [CADD] scores), along with pathway enrichment analysis of credible set variants (Oscanoa et al., 2020). A detailed description of these steps is provided in the **Supplementary information** (**paragraph 1.4**). Finally, the drugs/compounds that target genes mapped to likely causal variants were sourced from GeneCards, independent from their approved or investigational status. GeneCards is an online platform that gathers information from multiple databases including DrugBank, PharmaGKB, ClinicalTrials, DGIdb, the Human Metabolome Database, and Novoseek (Safran et al., 2010).

## 3. Results

### 3.1. Local patterns of genetic correlation between IR-related conditions and **neuropsychiatric disorders**

For each pair consisting of an IR-related condition and a neuropsychiatric disorder, bivariate local genetic correlation was evaluated in all genomic regions for which both phenotypes exhibited a univariate signal at p<1×10^-4^, resulting in a total of 2,251 tests. Of note, only 19.6% of the regions with significant local SNP-based heritability (h^2^) for both phenotypes showed a bivariate p<0.05, indicating that significant local h^2^_SNP_ is often present without any local correlation signal between neuropsychiatric and IR-related conditions. After FDR correction, moderate to high degrees of local genetic correlations (|r_g_|=0.21-1, p_FDR_<0.05) were identified for 20 of the 27 phenotype pairs examined, across 109 unique semi-independent genomic regions (see **Fig. 1** and **Table 2**). Noteworthy, local genetic correlations also emerged between IR-related conditions and neuropsychiatric disorders that had not shown significant global genetic correlations, namely AD, BD, and TS (Fanelli et al., 2022a). In total, 128 FDR-significant local genetic correlations were identified, of which 75 with a positive direction of the effect and 53 with a negative direction (**Table 2** ; detailed results are provided in **Table S1** ; see also **Fig. 1 b-c**). For 59 (46.1%) of the 128 local correlations, the 95% confidence intervals (CIs) for the explained variance included the value 1, consistent with a scenario where the local genetic signal for those phenotype pairs is entirely shared (**Table 2**). Interestingly, exclusively positive local genetic correlations were found between IR-related conditions and ADHD/MDD, while those detected between IR-related conditions and AN were all negative. No local genetic correlation was found between ASD and IR-related conditions. Conversely, a combination of positive and negative local genetic correlations was detected between all the other IR-related and neuropsychiatric conditions (**Fig. 1**, **Table 2**), of which all but MetS-schizophrenia had no previous evidence of global genetic overlap (see **Table 2**).

**Figure 1.**
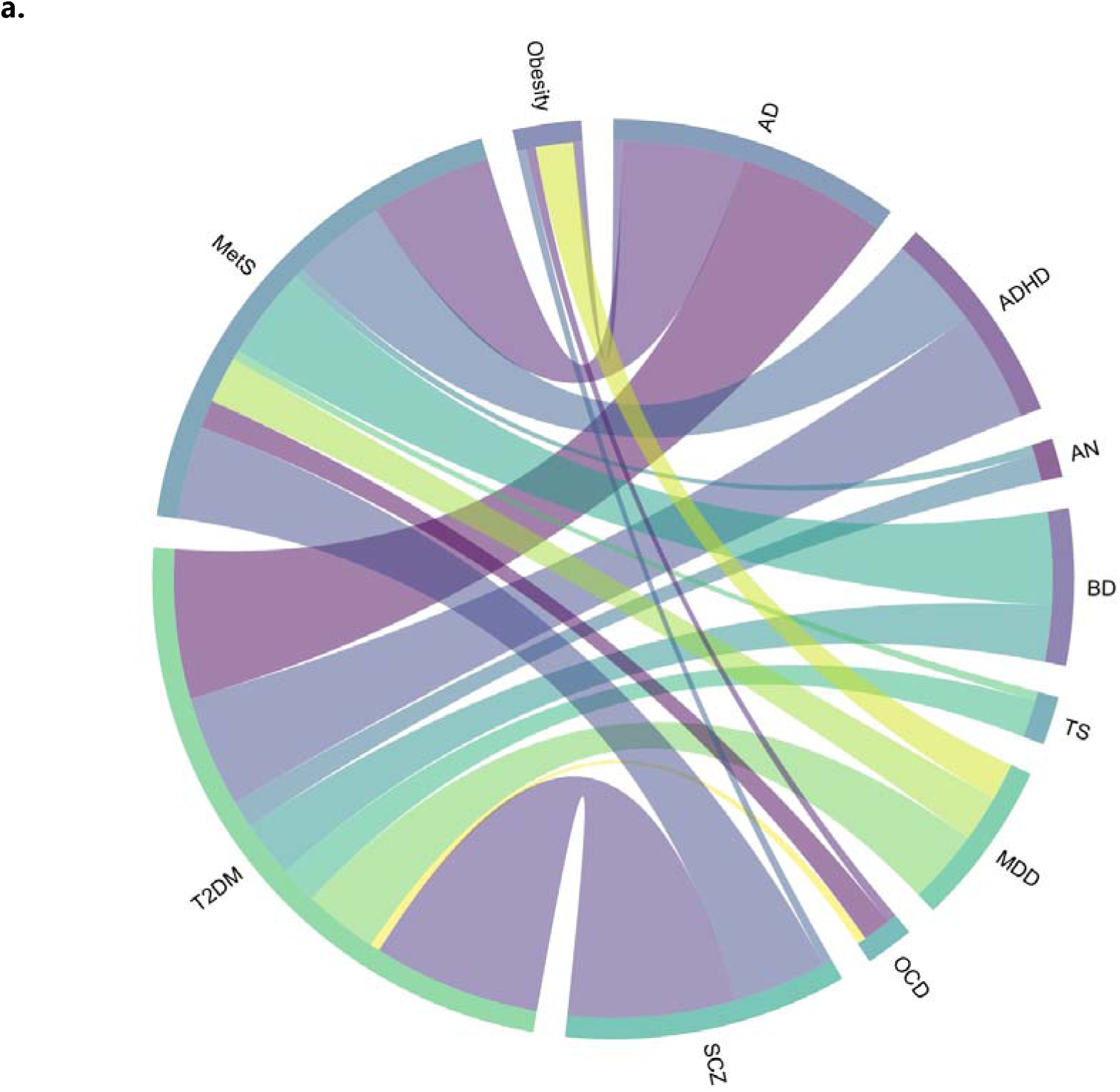
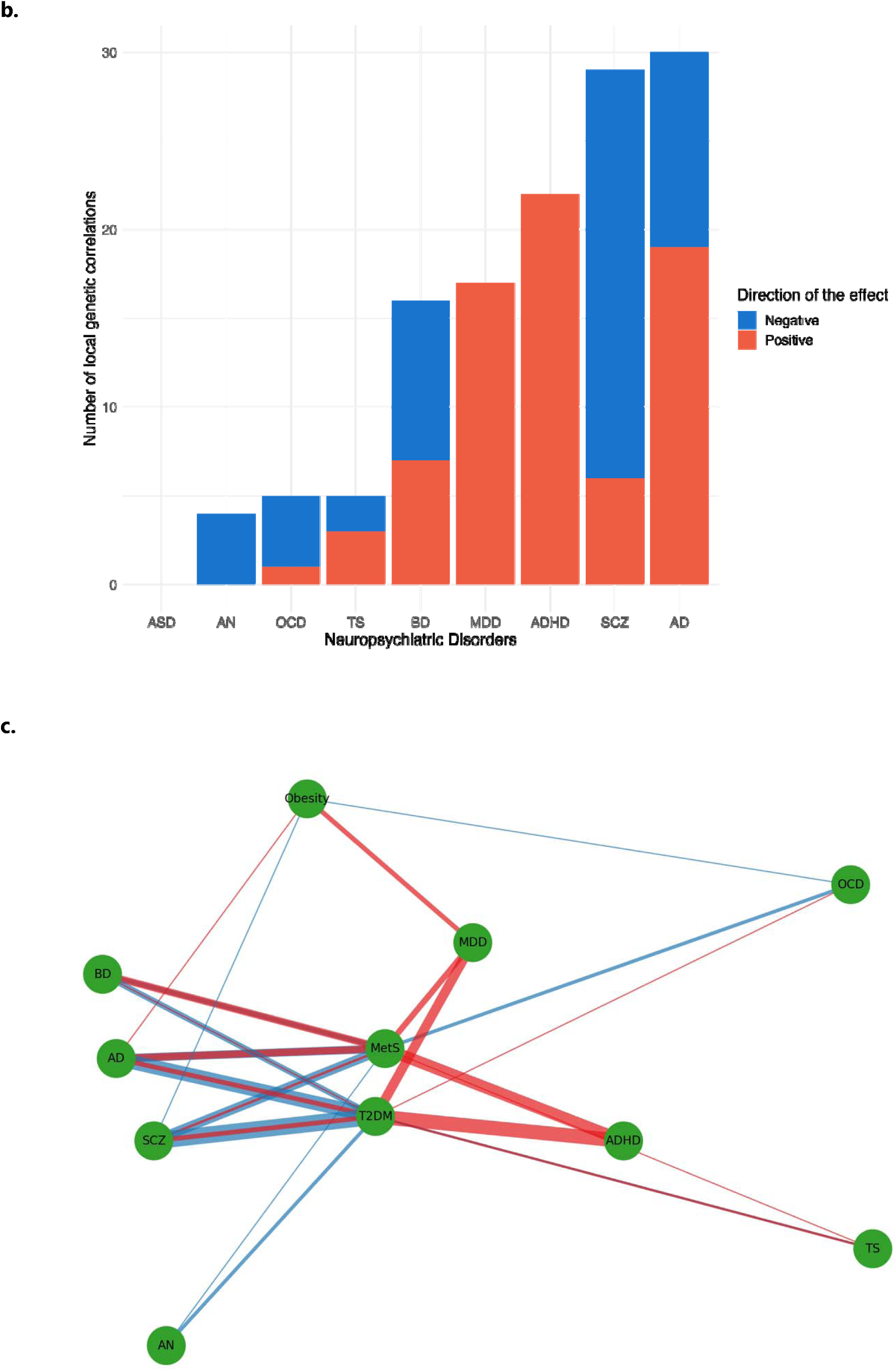
Local genetic correlations between insulin resistance-related conditions and neuropsychiatric disorders. a. Chord diagram representing the network of local genetic correlations between insulin resistance-related conditions and neuropsychiatric disorders. A higher width of a ribbon reflects a higher number of shared genetically correlated loci between two phenotypes, highlighting a substantial polygenic overlap and suggesting potential shared pathophysiological mechanisms between them. The colours of the ribbons are used purely for visual distinction and do not imply any additional significance or categorisation. b. Bar plot presenting the number of local genetic correlations identified between neuropsychiatric disorders and insulin resistance-related conditions. Each bar corresponds to a different neuropsychiatric disorder, segmented by the direction of effect of local genetic correlations, with blue indicating negative and red indicating positive local genetic correlations between neuropsychiatric disorders and insulin resistance-related conditions. The height of each bar reflects the quantity of local genetic correlations detected for each disorder. c. Network visualisation of local genetic correlations between a spectrum of neuropsychiatric disorders and insulin resistance-related conditions. Nodes represent distinct phenotypes for which local bivariate genetic correlations were evaluated. Edges connecting the nodes vary in width proportionally to the number of local genetic correlations identified between phenotype pairs. Edge colour denotes the direction of the genetic correlation estimate, with red indicating a positive correlation and blue indicating a negative correlation. Abbreviations: AD, Alzheimer’s disease; ADHD, attention-deficit/hyperactivity disorder; AN, anorexia nervosa; ASD, autism spectrum disorder; BD, bipolar disorder; MDD, major depressive disorder; MetS, metabolic syndrome, OCD, obsessive-compulsive disorder; T2DM, type 2 diabetes mellitus; SCZ, schizophrenia; TS, Tourette’s syndrome.

**Table 2 (on the next page).** Summary of local genetic correlations between neuropsychiatric disorders and insulin resistance-related conditions. For each neuropsychiatric disorder, the table presents the presence or absence of global genetic correlation with the specific insulin resistance-related condition, significant loci with h^2^_SNP_ for both phenotypes tested, and the directionality of local genetic correlations with T2DM, Metabolic Syndrome (MetS), and obesity. The table further provides the number of local genetic correlations with confidence intervals that include 1, indicating completely overlapping genetic influences at specific loci. Abbreviations: AD, Alzheimer’s disease; ADHD, attention-deficit/ hyperactivity disorder; AN, anorexia nervosa; ASD, autism spectrum disorder; BD, bipolar disorder; h^2^_SNP_, single-nucleotide polymorphism (SNP)-based heritability; MetS, metabolic syndrome; MDD, major depressive disorder; N, number of; OCD, obsessive-compulsive disorder; T2DM, type 2 diabetes mellitus; TS, Tourette’s syndrome. ^a^p<1×10-4; ^b^False discovery rate (FDR) at maximum q=0.05.

| Neuropsychiatric disorders | Insulin resistance-related conditions | Previous evidence of bivariate global genetic correlation | N loci with significant a local $h^2_{\text{SNP}}$ simultaneously for both phenotypes (% of loci tested) | N genetically correlated loci <sup>b</sup> (% of loci with significant <sup>a</sup> local $h^2_{\text{SNP}}$ for both phenotypes) | N positively correlated loci (% of correlated loci for the pair)<sup>b</sup> | N negatively correlated loci (% of correlated loci for the pair)<sup>b</sup> | N genetic correlations estimates whose confidence intervals included 1 (% of correlated loci for the pair)<sup>b</sup> |
| --- | --- | --- | --- | --- | --- | --- | --- |
| AD | MetS | NO | 306 (12.3%) | 13 (4.2%) | 6 (46.2%) | 7 (53.8%) | 2 (15.4%) |
| AD | Obesity | NO | 34 (1.4%) | 1 (2.9%) | 1 (100.0%) | 0 (0.0%) | 1 (100.0%) |
| AD | T2DM | NO | 214 (8.6%) | 16 (7.5%) | 12 (75.0%) | 4 (25.0%) | 3 (18.8%) |
| ADHD | MetS | YES | 94 (3.8%) | 10 (10.6%) | 10 (100.0%) | 0 (0.0%) | 9 (90.0%) |
| ADHD | Obesity | YES | 30 (1.2%) | 0 (0.0%) | 0 (0.0%) | 0 (0.0%) | 0 (0.0%) |
| ADHD | T2DM | YES | 88 (3.5%) | 12 (13.6%) | 12 (100.0%) | 0 (0.0%) | 7 (58.3%) |
| AN | MetS | YES | 65 (2.6%) | 1 (1.5%) | 0 (0.0%) | 1 (100.0%) | 1 (100.0%) |
| AN | Obesity | YES | 40 (1.6%) | 0 (0.0%) | 0 (0.0%) | 0 (0.0%) | 0 (0.0%) |
| AN | T2DM | YES | 73 (2.9%) | 3 (4.1%) | 0 (0.0%) | 3 (100.0%) | 2 (66.7%) |
| ASD | MetS | YES | 21 (0.8%) | 0 (0.0%) | 0 (0.0%) | 0 (0.0%) | 0 (0.0%) |
| ASD | Obesity | YES | 13 (0.5%) | 0 (0.0%) | 0 (0.0%) | 0 (0.0%) | 0 (0.0%) |
| ASD | T2DM | YES | 12 (0.5%) | 0 (0.0%) | 0 (0.0%) | 0 (0.0%) | 0 (0.0%) |
| BD | MetS | NO | 121 (4.8%) | 10 (8.3%) | 6 (60.0%) | 4 (40.0%) | 0 (0.0%) |
| BD | Obesity | NO | 53 (2.1%) | 0 (0.0%) | 0 (0.0%) | 0 (0.0%) | 0 (0.0%) |
| BD | T2DM | NO | 107 (4.3%) | 6 (5.6%) | 1 (16.7%) | 5 (83.3%) | 1 (16.7%) |
| MDD | MetS | YES | 95 (3.8%) | 5 (5.3%) | 5 (100.0%) | 0 (0.0%) | 3 (60.0%) |
| MDD | Obesity | YES | 45 (1.8%) | 4 (8.9%) | 4 (100.0%) | 0 (0.0%) | 3 (75.0%) |
| MDD | T2DM | YES | 83 (3.3%) | 8 (9.6%) | 8 (100.0%) | 0 (0.0%) | 4 (50.0%) |
| OCD | MetS | YES | 71 (2.8%) | 3 (4.2%) | 0 (0.0%) | 3 (100.0%) | 2 (66.7%) |
| OCD | Obesity | YES | 40 (1.6%) | 1 (2.5%) | 0 (0.0%) | 1 (100.0%) | 0 (0.0%) |
| OCD | T2DM | YES | 54 (2.2%) | 1 (1.9%) | 1 (100.0%) | 0 (0.0%) | 0 (0.0%) |
| Schizophrenia | MetS | YES | 200 (8.0%) | 10 (5.0%) | 2 (20.0%) | 8 (80.0%) | 6 (60.0%) |
| Schizophrenia | Obesity | NO | 89 (3.6%) | 1 (1.1%) | 0 (0.0%) | 1 (100.0%) | 1 (100.0%) |
| Schizophrenia | T2DM | NO | 172 (6.9%) | 18 (10.5%) | 4 (22.2%) | 14 (77.8%) | 10 (55.6%) |
| TS | MetS | NO | 57 (2.3%) | 1 (1.8%) | 1 (100.0%) | 0 (0.0%) | 1 (100.0%) |
| TS | Obesity | NO | 27 (1.1%) | 0 (0.0%) | 0 (0.0%) | 0 (0.0%) | 0 (0.0%) |
| TS | T2DM | NO | 47 (1.9%) | 4 (8.5%) | 2 (50.0%) | 2 (50.0%) | 3 (75.0%) |
| **TOTAL** |  |  | **2251** | **128 (5.7%)** | **75 (58.6%)** | **53 (41.4%)** | **59 (46.1%)** |

Furthermore, fifteen out of the 109 unique regions were associated with more than one phenotypic pair (**Table S1** ; we refer to these here as hotspots). The major hotspots showing significant bivariate local r_g_s between multiple phenotypic pairs were the chr2:59251997-60775066 (between T2DM-ADHD, MetS-AN, MetS-MDD), chr6:31320269-31427209 (MetS-AD, T2DM-AD, T2DM-schizophrenia), and chr16:29043178-31384210 genomic regions (MetS-schizophrenia, obesity-schizophrenia, T2DM-schizophrenia) (see **Table S1** and **Fig. S1**). Notably, 11.71% of the genetically correlated regions detected here (15/128) are located in the Major Histocompatibility Complex (MHC) region (chr6:28477797-33448354). All *r*_g_s detected in the MHC were between T2DM/MetS and either schizophrenia, AD, or BP, with prevalence of a negative direction of the effect (**Table S1**).

### 3.2. Genes underlying insulin resistance-neuropsychiatric multimorbidity

In the regions where we detected significant local genetic correlations, we identified a total of 1,455 distinct genes were identified through eQTL-based mapping, and 1,495 unique protein-coding genes through positional mapping across all phenotype pairs (**Table S2-3**). Notably, the pseudogene *CYP21A1P* was recurrently eQTL-mapped across multiple phenotype pairs (AD-T2DM, AD-MetS, BD-T2DM, schizophrenia-T2DM). In total, 140 genes were mapped for at least three phenotype pairs, indicating a potentially broader relevance in the genetic landscape of IR-neuropsychiatric multimorbidity (**Table S3**). Within this subset, 20 genes, all located within the MHC region, were involved in immune-inflammation and vesicle metabolism/trafficking (e.g., *HLA-B, MICA, C4A, C4B, AGER, BTNL2, HLA-DRA, HLA-DRB1, HLA-DQA1, PSMB8, HLA-DRB5,* and *FLOT1*), and four genes were involved in insulin signalling and secretion (i.e., *STX1A, FLOT1, MAPK3,* and *PHKG2*) (see **Table S3**).

### 3.3. Functional annotation of the identified regions

Considering the genes mapped to the regions showing local correlation, 411 gene sets were significantly enriched (**Table S4**). Immune-related pathways were prominently represented for multiple phenotype pairs (i.e., AD-MetS/T2DM, BD-T2DM, TS-T2DM, schizophrenia-T2DM). Other biological pathways related to oxygen transport, lipid metabolism (including omega-3 and omega-6 polyunsaturated fatty acid levels (PUFAs)), embryonic/placental development, insulin receptor/phosphoinositide 3-kinase (PI3K), and vesicular function/secretion were enriched across different phenotype pairs (**Table S4**). Pharmacogenomic markers, notably genes genome-wide associated with response to metformin (i.e., *STX1B, STX4, ZNF668*), were enriched in regions shared between schizophrenia and MetS, obesity, and T2DM (**Table S5**).

In a more granular examination, we also evaluated enrichment of life-stage-specific expression profiles for genes mapped to the genetically correlated regions (**Tables S7-8**). Specifically, regions correlated between schizophrenia and obesity featured genes upregulated at 19 weeks post-conception. Conversely, regions associated with the schizophrenia-MetS pair exhibited a distinct pattern, with genes showing downregulation in brain samples from individuals at age 11. Furthermore, regions of overlap between OCD and MetS held genes upregulated in early adulthood brain tissues, while the genes in the overlapping regions marking the OCD-obesity pair exhibited gene downregulation in late childhood.

Detailed results for gene set analysis, spatio-temporal expression specificity of the mapped genes, and druggable gene annotations are reported in **Table S4-S10**.

### 3.4. Shared causal variants between insulin resistance-related conditions and neuropsychiatric disorders

Of the 128 regions identified with local r_g_, colocalisation analyses successfully pinpointed the likely causal variants driving this association in 10 regions (see **Table S11-12**). For comprehensive functional annotations of 95% credible set variants within these 10 regions see **Tables S13-S23**.

Notably, one region on chromosome 4 and two on chromosome 6 showed the highest posterior probability for colocalisation, linking schizophrenia with MetS and AD with T2DM, respectively (**Table S11-12**, **Fig. S2-4**). The schizophrenia-MetS relationship implicated the rs13107325 variant in the *SLC39A8* gene, which modulates the activity of the miRNA hsa-miR-374b-5p (**Tables S12-S14**). For the AD-T2DM pair, the likely causal variants were rs9271608 and rs9275599, mapped to the *HLA-DRB1* and *MTCO3P1* genes, respectively. According to GeneCards, *HLA-DRB1* is targeted by immunosuppressive and anti-inflammatory drugs (e.g., azathioprine, lapatinib, interferons-β, and acetylsalicylic acid), as well as by statins and psychotropic drugs (e.g., carbamazepine, clozapine, and lamotrigine) (**Table S23**).

Further seven regions had good support for colocalisation (**Supplementary information, paragraph 1.3**); these regions showed local genetic correlations for the AD-T2DM, MDD-T2DM, BD-MetS, and schizophrenia-MetS pairs (**Table S11-12**). Most of the identified variants were observed within or near genes pivotal to immune function, vesicle/small molecules trafficking, lipid metabolism, organ development, retinoic acid signalling, and DNA repair/apoptosis (**Tables S17-18**). They often had high CADD PHRED scores, suggesting highly deleterious effects (**Tables S13**). Genes mapping to these variants, like the *HLA-DQB1* and *FADS1/2* genes, are targeted by existing drugs and supplements, such as antihypertensive drugs, omega-3/6 PUFAs, and vitamin A (**Table S23**).

## 4. Discussion

In this study, we examined the genetic relationship between IR-related conditions - specifically, obesity, T2DM, and MetS - and nine neuropsychiatric disorders by investigating the pairwise patterns of local genetic correlation across the genome, while exploring the specific genetic factors and biological mechanisms underlying their multimorbidity. The results presented here offer novel insights into the shared genetic aetiology between these phenotypes, unveiling a complex pattern of both positive and negative local genetic correlations. For the first time, we demonstrated that even in the absence of global genetic correlations, significant local correlations exist (i.e., between AD, BD, TS and IR-related conditions), expanding the results of previous studies (Fanelli et al., 2022a; Hubel et al., 2019), with important implications on the pathophysiology of these disorders and on new avenues for targeted therapeutic interventions addressing the issue of IR-psychiatric multimorbidity. We identified 128 local genetic correlations across 109 unique genomic regions. Notably, the MHC region emerged as a particularly significant contributor in terms of shared genetic signal, as confirmed by enrichment in biological pathways related to immune function. Genes mapped to the genetically correlated regions also showed enrichment in pathways involved in lipid metabolism, insulin signalling, and vesicular function, offering new insights into the biological mechanisms at play.

Regarding the directions of the detected genetic correlations, we observed exclusively positive local genetic correlations for ADHD and MDD with IR-related conditions, indicating synergistic genetic effects that predispose to both neuropsychiatric symptoms and IR-related conditions. Our enrichment analyses of the genes mapped to these regions suggest that the genetic overlap might be mediated by genes involved in extracellular matrix organisation, vesicle trafficking, and oxygen transport/oxidative processes. These pathways are crucial in both brain function and metabolic regulation (Dityatev et al., 2010; Rossetti et al., 2020; Zou et al., 2020). Extracellular matrix molecules are implicated in synaptic plasticity and homeostasis (Dityatev et al., 2010) and may also influence tissue insulin sensitivity (Williams et al., 2015). Similarly, vesicle trafficking, integral to synaptic function and neurotransmission, could be a nexus point where neuronal communication and insulin signalling intersect, contributing to the multimorbidity of the conditions (Zou et al., 2020). Conversely, we detected exclusively negative correlations between IR-related conditions and AN. These results align with the distinct phenotypic characteristics of AN, including increased insulin sensitivity and metabolic alterations related to undernutrition, which differ markedly from other neuropsychiatric disorders (Duriez et al., 2019; Ilyas et al., 2019).

While phenotypic overlap of AD and BD with IR-related conditions has been reported frequently (e.g., Santiago and Potashkin (2021); Wimberley et al. (2022)), previous genetic analyses did not find global genetic correlations between these phenotypes (Fanelli et al., 2022a), potentially due to the averaging effect of global analyses. Our study, which is the first to report significant local genetic correlations between AD, BD, TS and IR-related conditions, suggests that positive and negative local correlations may regularly neutralise each other in global correlation analyses, a phenomenon observed in other recent studies (Arenella et al., 2023; Fernandes et al., 2023), and should be taken into account for future research. These heterogeneous patterns of genetic overlap could point towards aetiologically distinct subgroups, which could be examined in greater detail using deep phenotyping and functional validation. Such analyses could bring us closer towards precision medicine, offering the potential of personalising healthcare and increase treatment success (Feczko and Fair, 2020).

Some genomic regions (15 out of 109) showed significant correlations for multiple phenotype pairs, implying a potentially more prominent and ubiquitous role in the IR-neuropsychiatric multimorbidity. Among the recurring regions, chr2:59251997-60775066, mapping to the *BCL11A* gene, was implicated in the correlation of T2DM with ADHD, MetS and AN, and of MetS with MDD. *BCL11A* codes for a transcription factor with a central role in B cells and haematopoiesis as well as in neuronal development, regulating processes such as neurogenesis/axonogenesis, and neuronal migration (Bauer and Orkin, 2015; Dias et al., 2016). *BCL11A* variants have also been associated with neurodevelopmental disorders and impaired cognition, as well as with IR in *in vivo* and *in vitro* studies (Dias et al., 2016; Jonsson et al., 2013; Wiegreffe et al., 2022). Among other genes that were mapped across at least three phenotypic pairs, some (i.e. *STX1A, FLOT1, MAPK3*, and *PHKG2*) were found to have pivotal roles in insulin signalling and secretion (Bagge et al., 2013; Jager et al., 2011; van de Vondervoort et al., 2016). These findings strengthen a molecular basis for the connection between altered insulin function and neuropsychiatric disorders (Fanelli et al., 2022a; van de Vondervoort et al., 2016)(NR Mota, in preparation), which has been tied to cognitive deficits, anhedonia, and reward processing alterations (Fanelli et al., 2022b; Fanelli and Serretti, 2022; Possidente et al., 2023).

Over 11% of the correlated genomic regions were located within the MHC region (chr6:28477797-33448354), where extensive pleiotropy has been demonstrated previously (Watanabe et al., 2019; Werme et al., 2022). This region is renowned for its high gene density, polymorphism, and involvement in immune-inflammatory responses (Matzaraki et al., 2017). The influence of the MHC region extends beyond autoimmune and infectious diseases susceptibility, being also associated with neuropsychiatric disorders, such as ASD, schizophrenia, and BD, underscoring its broad impact on health (Tamouza et al., 2021). Our findings point to a plausible genetic link between IR-related metabolic dysfunction, immune-inflammatory dysfunction, and neuropsychiatric disorders. This is consistent with previous findings indicating that central and peripheral inflammation may mediate the link between IR and neuropsychiatric conditions (Chan et al., 2019; Viardot et al., 2012). Inflammation may also impair brain insulin signalling, leading to neurobehavioural damage (Gong et al., 2019). Notably, most of the local genetic correlations identified within the MHC region showed a negative direction of effect. We cannot provide a clear explanation of this finding, but it may lie in the intricate balance of pro-inflammatory and anti-inflammatory factors in immune response, in which MHC genes play a critical role (Tamouza et al., 2021). The differential expression of genes in the MHC region across different tissues and lifespan could also contribute to this observation (Shen and Zhang, 2021). Experimental validation of these findings will therefore be necessary to determine the exact functional implication of the observed genetic associations.

Relatedly, our study identified multiple genes related to the human leukocyte antigen (HLA) system, innate immunity, and immunomodulation (i.e., *HLA-B, HLA-DRA, HLA-DRB1, HLA-DQA1, HLA-DRB5, MICA, C4A, C4B, AGER, PSMB8,* and *BTNL2*), supporting their possible influence on IR-neuropsychiatric multimorbidity. Of note, immunomodulatory drugs (e.g., non-steroidal anti-inflammatory drugs and monoclonal antibodies) have shown some efficacy as add-on treatments in psychoses and MDD, and might have higher efficacy in people with IR-neuropsychiatric multimorbidity (Drevets et al., 2022; Jeppesen et al., 2020). Another gene recurrently mapped across various phenotype pairs was the *CYP21A1P* pseudogene, located within the MHC region. Intergenic recombination of *CYP21A1P* leads to altered glucocorticoid and androgen production (Carvalho et al., 2021); glucocorticoids possess anti-inflammatory/immunosuppressive effects, and regulate glucose metabolism and the body’s stress response (Balsevich et al., 2019). Specifically, glucocorticoids counteract insulin by decreasing peripheral glucose uptake and stimulating hepatic gluconeogenesis, leading to IR under conditions of excessive release, such as in chronic stress (Fichna and Fichna, 2017). Prolonged exposure to glucocorticoids can induce neurotoxic effects, possibly involved in the development of psychiatric disorders (Chiba et al., 2012; Ding et al., 2022). Furthermore, glucocorticoids modulate the serotonergic system, which is strongly implicated in psychiatric disorders and insulin signalling (Betari et al., 2021; Prouty et al., 2019). Interestingly, gene set enrichments within correlated regions between schizophrenia and IR-related conditions were related to the response to metformin, a frontline oral medication for T2DM. This implies a potential overlap in therapeutic targets between schizophrenia and T2DM, which could lead to a reassessment of treatment strategies for these patients. Previous randomised-controlled trials (RCTs) confirmed the efficacy of metformin in contrasting antipsychotic-induced metabolic side effects in individuals with psychoses (Agarwal et al., 2021; de Silva et al., 2016), while improving psychiatric and cognitive symptoms in the same population (Battini et al., 2023).

Another finding of this study was the identification of significant colocalisation signals. Among the 128 regions demonstrating local genetic correlation, 10 key regions were prioritised for their high posterior probabilities of housing the same shared causal variants between IR-related conditions and neuropsychiatric disorders. This was instrumental for further elucidating shared pathophysiological mechanisms and novel potential drug targets for IR-neuropsychiatric multimorbidity (Belyaeva et al., 2021; Karki et al., 2017). The two most likely shared causal variants were located in the chr4:102544804-104384534 and chr6:32586785-32629239/chr6:32682214-32897998 regions, suggesting novel cross-links between schizophrenia and MetS, and AD and T2DM, respectively. The identified shared causal variant (rs13107325) between schizophrenia and MetS maps to the *SLC39A8* gene, encoding the ZIP8 metal cation transporter. Previous studies demonstrated its association with altered brain manganese levels and protein complexity in schizophrenia, brain morphology and dendritic spine density, as well as a broader impact on various conditions, including developmental, neuropsychiatric and cardio-metabolic diseases/traits (Hermann et al., 2021; Li et al., 2022; Mealer et al., 2020; Nebert and Liu, 2019). Our findings also highlight SLC39A8’s potential as a therapeutic target via zinc chloride/sulphate (Wishart et al., 2018). Interestingly, RCTs have shown beneficial effects of zinc sulphate in reducing symptoms of ADHD, MDD and SCZ (Behrouzian et al., 2022; Bilici et al., 2004; Salari et al., 2015), as well as improving glucose handling in prediabetes (Islam et al., 2016). In the AD-T2DM context, the rs9271608 variant mapping to the *HLA-DRB1* gene presented compelling causal candidacy, pointing to the potential for immunosuppressive drugs such as azathioprine, lapatinib, and interferons-β to influence AD-T2DM manifestations. The administration of intranasal treatment with interferon-β was shown to improve anxious/depressive-like behaviours by modulating microglia polarisation in AD rat models (Farhangian et al., 2023). Of note, the rs9271608 also shows broad biological relevance as it is active as a promoter across numerous cell types and tissues, including various immune and neuronal progenitors (Zerbino et al., 2015). The remaining regions of notable colocalisation underpin associations between AD and T2DM, MDD and T2DM, BD and MetS, and schizophrenia and MetS, hinting at potential targetable mechanisms for current drugs and supplements, including antihypertensive drugs, omega-3/6 PUFAs, vitamin A (Wishart et al., 2018). Several antihypertensive drugs have been associated with a reduced risk of depression (Kessing et al., 2020), and omega-3 PUFAs showed beneficial effects on depression symptoms in a meta-analysis of RCTs (Liao et al., 2019). Genes associated with omega-3/omega-6 PUFAs were enriched when considering the regions showing correlation between BD and MetS, in line with their relevance in the multimorbidity. Finally, vitamin A inhibits amyloid β protein deposition, tau phosphorylation, neuronal degeneration and improves spatial learning and memory in AD mouse models (Ono and Yamada, 2012). It is worth noting that a significant local genetic correlation without detectable colocalisation does not necessarily mean that there are no shared causal variants; this may reflect limitations in the power of the colocalisation analysis, particularly in scenarios with complex patterns of associations, which are often observed in highly polygenic traits (Werme et al., 2022).

Our study should be viewed considering some limitations. Although our study pinpoints shared causal variants and proposes biological mechanisms through which shared genetic regions might impact mental and metabolic health, the functional interpretation of these findings remains largely speculative, requiring *in vitro* validation or studies in animal models. The high LD in the MHC region may have led to spurious pleiotropy, not necessarily implying the presence of the same shared causal SNPs (Lee et al., 2021). Rare genetic variants were not considered, and population-specific effects may not be adequately captured by our analyses, which were limited to European ancestry. While the available GWAS summary statistics were generally obtained in samples of adequate size for this kind of study, the GWAS summary statistics for OCD were based on a relatively small sample size, potentially influencing the number of significant local genetic correlations detected by LAVA.

### 4.1. Conclusions

In conclusion, our study provides a pivotal contribution to the understanding of the genetic underpinnings of neuropsychiatric and IR-related conditions, challenging traditional notions of their separate pathophysiology. Our result support a more integrated disease model, and the need to move beyond the conventional view of distinct aetiologies. The implications of our findings extend to clinical practice, emphasising the need for a holistic approach in the screening and management of IR-neuropsychiatric multimorbidity. For example, the importance of lifestyle interventions for both metabolic and psychiatric health, and of developing pharmacological treatments that target both conditions. The discovery of shared causal variants, particularly in genes like *SLC39A8* and *HLA-DRB1*, opens new avenues for targeted therapeutic interventions. The convergence of genetic findings on mechanisms related to immune-inflammation, insulin signalling, lipid metabolism, vesicle trafficking, among others, provides a compelling direction for future research. Overall, our study not only unveils the shared genetic landscape of neuropsychiatric and IR-related conditions, but also establishes a foundation for integrated research and treatment approaches, contributing to a paradigm shift towards comprehensive care strategies that address the issue of multimorbidity.

### URLs

Local Analysis of [co]Variant Association (LAVA): https://github.com/josefin-werme/LAVA; LD SCore (LDSC) https://github.com/bulik/ldsc; biomaRt: https://bioconductor.org/packages/release/bioc/html/biomaRt.html; LiftOver: https://genome.ucsc.edu/cgi-bin/hgLiftOver; loci2path: https://www.bioconductor.org/packages/release/bioc/html/loci2path.html; coloc & SuSiE: https://github.com/chr1swallace/coloc; SNPnexus: https://www.snp-nexus.org; Functional Mapping and Annotation of Genome-Wide Association Studies (FUMA): http://fuma.ctglab.nl.

### Author contributions

GF: Conceptualisation, Data Curation, Formal analysis, Methodology, Software, Validation, Visualisation, Investigation, Writing—original draft preparation; BF: Supervision, Conceptualisation, Project administration, Funding acquisition, Resources, Writing—review and editing; CF: Supervision, Funding acquisition, Writing—review and editing; JW: Data Curation, Methodology, Software, Writing—review and editing; IE: Software, Writing—review and editing; WDW: Data Curation, Software, Writing—review and editing; GP: Writing— review and editing; IHR: Writing—review and editing; LMR: Data Curation, Writing—review and editing; VvG: Writing—review and editing; WJJ: Writing—review and editing; SJBV: Writing— review and editing; KAA: Writing—review and editing; AM: Writing—review and editing; JH: Writing—review and editing; TW: Writing—review and editing; SD: Writing— review and editing; AF: Writing—review and editing; CB: Writing—review and editing; FFA: Writing—review and editing; SJM: Writing—review and editing; SB: Writing—review and editing; SM: Writing—review and editing; JSS: Writing—review and editing; MA: Conceptualisation; Writing—review and editing; AS: Writing—review and editing, Funding acquisition; NRM: Conceptualisation; Writing—review and editing; JB: Supervision, Conceptualisation, Writing—review and editing, Funding acquisition, Resources.

## Data Availability

All data produced in the present work are contained in the manuscript

## Acknowledgements

This project has received funding from the European Union’s Horizon 2020 research and innovation programme under grant agreement No 847879 (PRIME, Prevention and Remediation of Insulin Multimorbidity in Europe). This publication is part of the project ‘No labels needed: understanding psychiatric disorders based on genetic traits’ (with project number 09150161910091) of the research program Veni, which is partly financed by the Dutch Research Council (NWO) Health Research and Development (ZonMW). The project also received relevant funding from the Netherlands Organization for Scientific Research (NWO) for the GUTS project (grant 024.005.011). Research reported in this publication was supported by the National Institute Of Mental Health of the National Institutes of Health under Award Number R01MH124851. The content is solely the responsibility of the authors and does not necessarily represent the official views of any of the funders. JSS is partially supported by ICREA under the ICREA Academia programme. FFA and SJM thank CERCA Programme/Generalitat de Catalunya for institutional support. This work was carried out on the Dutch national e-infrastructure with the support of SURF Cooperative.

