## Supplementary information for "Local patterns of genetic sharing challenge the boundaries between neuropsychiatric and insulin resistance-related conditions"

### 1.1. **Local genetic correlation analyses**

First, we generated 2,495 semi-independent genomic regions of ~1Mb by recursively splitting chromosomes while minimising linkage disequilibrium (LD) between them, following the approach reported by Werme et al. (2022). For this process, we used the 1000 Genomes Project, phase 3 (European ancestry, GRCh37/hg19 build) as our reference genome and the Local Analysis of [co]Variant Association (LAVA) partitioning algorithm (<https://github.com/cadeleeuw/lava-partitioning>; de Leeuw (2021)). Second, univariate analyses were conducted using LAVA to assess single-nucleotide polymorphism (SNP)-based heritability (h^2^_SNP_) at the level of each individual genomic region for each individual IR-related and neuropsychiatric phenotype. We selected only those genomic regions exhibiting significant local h^2^_SNP_ values (univariate p-value<1x10^-4^) for the subsequent bivariate analyses. Third, local genetic correlations were estimated between IR-related and neuropsychiatric phenotype pairs at each individual genomic region passing univariate analyses. To account for any potential sample overlap in the input summary statistics, we suppled the intercepts from the bivariate cross-trait LD Score (LDSC) analyses to LAVA as these represent an estimate of the sampling correlation between the data sets (Bulik-Sullivan et al., 2015).

### 1.2. **Functional annotation of genetically correlated regions**

For analysing the gene expression profiles, data from the Genotype-Tissue Expression (GTEx) project (Consortium, 2015) and the BrainSpan Atlas of the Human Brain were used, the latter including information from 11 general developmental stages and 29 distinct ages of brain samples (Kang et al., 2011; Miller et al., 2014). This analysis was complemented by an assessment of the tissue specificity of these genes, through the computation of the tissue-specificity index (tau) and identification of differentially expressed genes (DEGs) across tissues, offering further insight into their potential functional significance (Watanabe et al., 2017).

To investigate the involvement of the mapped genes in various biological pathways and processes, gene set enrichment analyses were carried out using hypergeometric tests. Enrichment analyses were conducted considering hallmark gene sets, which represent well-defined biological states or processes; ontology gene sets, which comprise genes annotated by the same ontology term; immunologic signature gene sets, representing cellular states and perturbations within the immune system; canonical pathway gene sets, which are collections of genes that work together to perform a specific biological function and are well-documented in scientific literature; and regulatory target gene sets, based on microRNA seed sequences and predicted transcription factor binding sites (Watanabe et al., 2017). Ontology gene sets were sourced from the Gene Ontology (GO) database, which collects predefined gene sets related to biological processes, cellular components, and molecular functions (The Gene Ontology, 2019). Canonical pathway gene sets were sourced from the BioCarta, Kyoto Encyclopedia of Genes and Genomes (KEGG) and Reactome databases (Fabregat et al., 2016; Kanehisa and Goto, 2000; Nishimura, 2001).

### 1.3. **Colocalisation analyses**

The coloc method utilises a Bayesian framework to estimate the posterior probabilities (PPs) of five different hypotheses (i.e., H0: no association with either trait; H1: association with trait 1, not with trait 2; H2: association with trait 2, not with trait 1; H3: association with trait 1 and trait 2, two independent SNPs; H4: association with trait 1 and trait 2, one shared SNP) representing the shared or distinct causal variants between the two phenotypes (i.e., insulin resistance-related conditions and neuropsychiatric disorders). By integrating the summary statistics from two respective GWASs, the method calculates the PPs for each hypothesis and assesses the likelihood of colocalisation. A high PP for hypothesis H4 (PP.H4) indicates strong evidence for colocalisation, whereas a low PPH4 suggests distinct causal variants. These PPs are computed by taking into account the SNP-trait association statistics (e.g., p-values or Z-scores) and the LD structure among the SNPs. The LD structure of the correlated genomic regions was determined using PLINK 1.9 (Purcell et al., 2007). The coloc analysis was executed using the coloc.abf() function with default parameters (Giambartolomei et al., 2014). Genomic loci harbouring plausible causal variants affecting both traits were identified based on the PPs resulting from the colocalisation analyses (Huang et al., 2020; Schmiedel et al., 2021). In particular, genetically correlated loci were selected only if the cumulative PPs for hypotheses H3 and H4 exceeded 0.8, thereby ensuring a substantial degree of confidence that the identified loci host at least one causal variant for either or both traits under investigation. Moreover, we imposed a ratio constraint wherein the PP.H4 needed to be at least five-fold greater than PP.H3. This second criterion enriched our selection for genomic loci wherein the evidence supports a shared causal variant influencing both traits, as opposed to distinct causal variants for each trait (Huang et al., 2020; Schmiedel et al., 2021). After identifying genomic loci that met our pre-established condition (i.e., PP.H3+PP.H4≥0.8-PP.H4/PP.H3≥5), we pinpointed the most likely causal variant for the shared signal based on either a PP.H4 of the individual SNP (SNP.PP.H4)≥0.5 or the calculation of credible sets of variants, which are pivotal for narrowing down potential causal variants. Using Bayesian methods, we derived the minimal set of SNPs that may encompass all causal variants with 95% probability (α). To construct our α-credible sets, we ranked the SNPs within each locus according to their SNP.PP.H4 values, from highest to lowest. We then computed the cumulative sum of SNP.PP.H4s until it equalled or exceeded α. This procedure ensures that the resulting set of SNPs is the smallest subset accounting for at least 95% of the PP of containing the causal shared variant (Schaid et al., 2018). By applying this rigorous method, we were able to reduce the list of potentially causal variants, while retaining high confidence in the likelihood of these sets containing the true causal variants. Subsequent to the identification of the most likely causal variants, we employed LocusZoom to visually represent the genomic context and LD patterns surrounding these GWAS associated variants. This visualisation was conducted separately for each phenotype and then combined to illustrate the patterns for both phenotypes collectively (Pruim et al., 2010).

### 1.4. **Functional annotation of 95% credible sets of shared causal variants**

Our annotation process involved an in-depth analysis using SNPnexus to characterise the functional significance of likely causal variants. Combined Annotation Dependent Depletion (CADD) scores integrates multiple annotations into one metric by contrasting variants that survived natural selection with simulated mutations, thereby predicting the functional impact of variants on protein-coding sequences (Kircher et al., 2014). For the purpose of our analysis, a CADD-PHRED score≥20 served as our threshold for identifying potentially deleterious variants falling within the top 1% of most deleterious substitutions in the human genome, while a CADD-PHRED≥10, which corresponds to the top 10% most deleterious substitutions, was also used as a less stringent threshold. The PHRED-like score is a transformation of the raw score that ranks the variant relative to all possible 8.6 billion variants in the human genome. Complementing the CADD scores, we analysed the transcription factor binding sites (TFBSs) related to our most likely causal SNPs (Wingender et al., 2001). TFBSs are specific DNA regions where transcription factors bind, influencing the regulation of gene expression. Moreover, the analysis incorporated information from miRBase, a comprehensive database of miRNA sequences and annotation. MicroRNAs (miRNAs) are pivotal non-coding RNAs that influence gene expression. We also considered the localisation of variants in predicted CpG (cytosine-phosphate-guanine) islands, which are genomic regions rich in cytosine-guanine dinucleotides, often linked to gene promoters. Other databases like TarBase and TargetScan also contributed to our analysis by providing experimentally validated data on miRNA targets and miRNA biological target predictions, respectively (Lewis et al., 2005; Papadopoulos et al., 2009). Other regulatory elements in the human genome were annotated using RegBuild, a resource from Ensembl (Zerbino et al., 2015). Additionally, the Roadmap Epigenomics Project data were considered to enrich our analysis by providing comprehensive epigenetic maps indicative of chromatin states that further elucidate potential functional elements in the regions of interest (Romanoski et al., 2015). The annotation process utilised the GRCh37/hg19 assembly of the human genome from Ensembl as the reference, supplementing it with additional databases including RefSeq, UCSC, GENCODE, and VEGA to ensure a thorough annotation (Oscanoa et al., 2020). Additionally, within the SNPnexus platform, pathway analysis was conducted focusing on 95% credible set variants. Within each query set, SNPnexus links genes to associated biological Reactome pathways. This process involves mapping variants in each credible set to their respective genes. Subsequently, the number of genes in the Reactome universe, as well as the number of genes in each pathway was determined. These parameters serve as inputs for Fisher’s Exact tests, which assess whether the number of affected genes in each set significantly deviates from what would be expected by random chance for each Reactome pathway. This approach allows us to identify pathways that are over-represented within credible set variants, providing further insights into potential causal mechanisms underlying neuropsychiatric-insulin resistance multimorbidity (Oscanoa et al., 2020).
