## Supplementary figures for "Local patterns of genetic sharing challenge the boundaries between neuropsychiatric and insulin resistance-related conditions"

Supplementary Figure 1. Network plot depicting local genetic correlations between schizophrenia (SCZ) and type 2 diabetes mellitus (T2DM), obesity, and metabolic syndrome (MetS) for the genomic region chr16:29043178-31384210, partly mapping to the Major Histocompatibility Complex (MHC) region. Each node corresponds to one of the phenotypes, and the connecting lines represent the strength and direction of the genetic correlation, with the width of the lines proportional to the absolute value of the local genetic correlation estimate (|rho|). The gradient from red to blue indicates the direction of the local genetic correlation estimates, with red signifying a positive correlation and blue a negative correlation. This visualisation underscores the complex genetic interrelations in the MHC region that may underlie the comorbidity between SCZ and T2DM, obesity, and MetS.

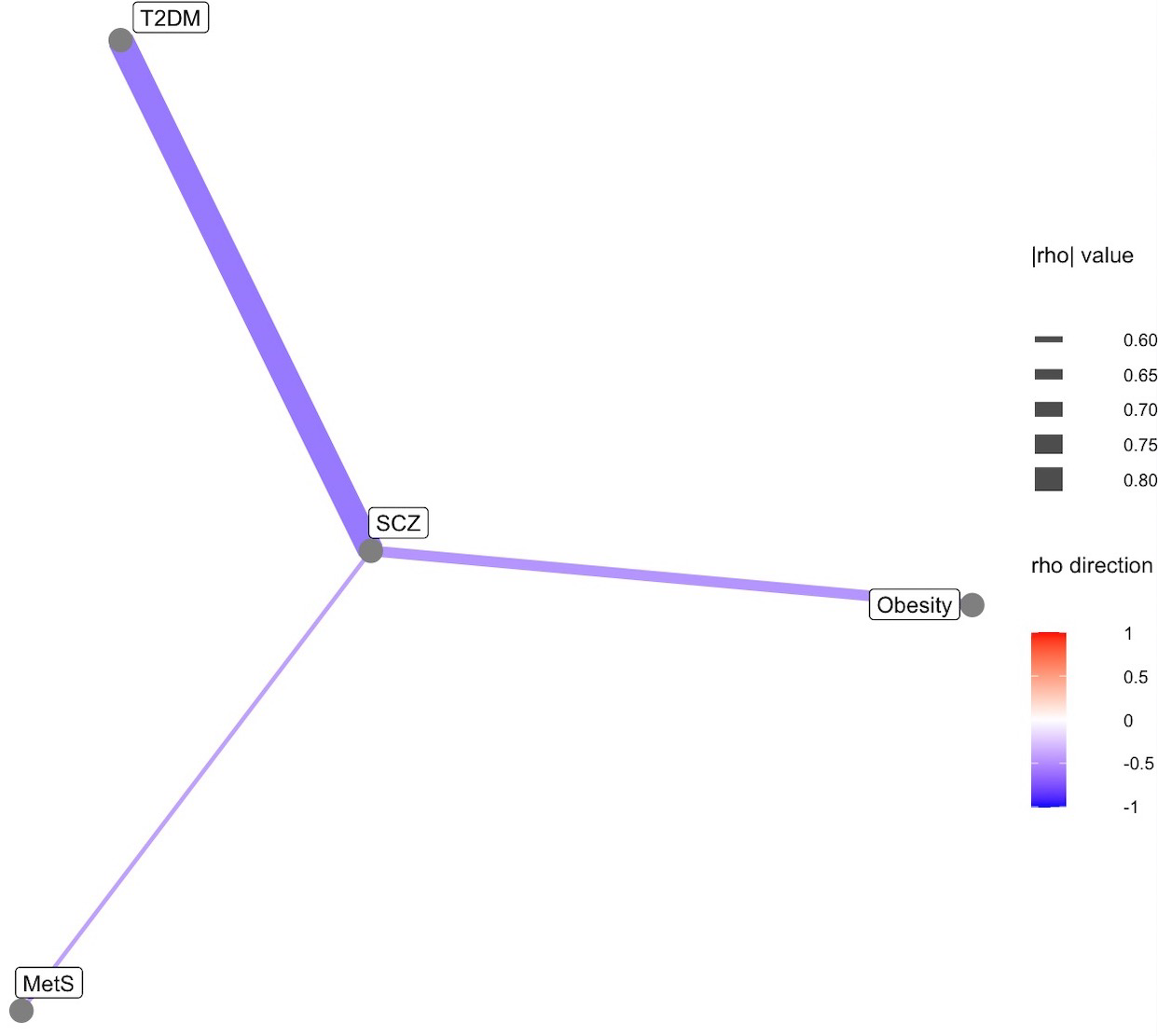

Supplementary Figure 2. Bivariate LocusPlot depicting the genome-wide association signals across the chromosomal region 4:102544804-104384534 for schizophrenia (SCZ) and metabolic syndrome (MetS). The plot on the left illustrates the −log10(p)-values from the GWAS on MetS (x-axis) versus SCZ (y-axis), with each point representing a single nucleotide polymorphism (SNP). The colour of each SNP denotes the linkage disequilibrium (LD) with the lead SNP rs13107325, as indicated by the colour scale. The plots on the right focus on the physical position along chromosome 4 (x-axis) against the −log10(p)-values (y-axis) of association in the GWAS on SCZ and MetS. The lead SNP rs13107325, which has a high posterior probability (PP) of being a shared causal variant, is prominently highlighted in both plots on the right. This indicates a genomic region of interest that may harbour pleiotropic causal effects, potentially implicating this locus in the comorbidity of SCZ and MetS.

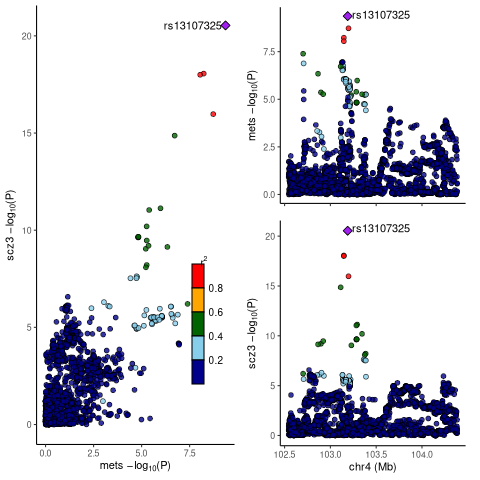

Supplementary Figure 3. Bivariate LocusPlot depicting the genome-wide association signals across the chromosomal region 6:32586785-32629239 for Alzheimer’s disease (AD) and type 2 diabetes mellitus (T2DM). The plot on the left illustrates the −log10(*p*)-values from the GWAS on T2DM (x-axis) versus AD (y-axis), with each point representing a single nucleotide polymorphism (SNP). The colour of each SNP indicates the linkage disequilibrium (LD) with the lead SNP, as reflected by the colour scale. The plots on the right focus on the physical position along chromosome 6 (x-axis) against the −log10(*p*)-values (y-axis) of association in the GWAS on AD and T2DM. The lead SNP, which has a high posterior probability (PP) of being a shared causal variant, is prominently highlighted in both plots on the right. This indicates a genomic region of interest that may harbour pleiotropic causal effects, potentially implicating this locus in the comorbidity of AD and T2DM.

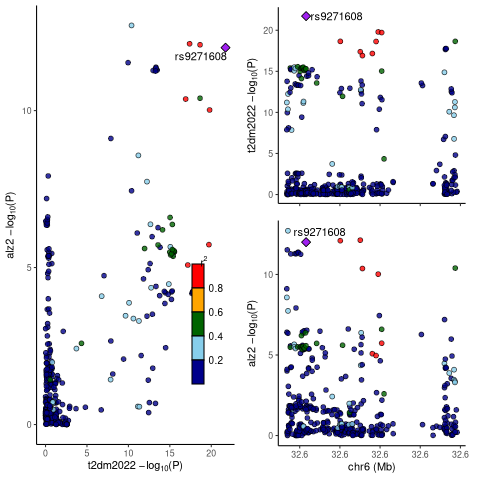

Supplementary Figure 4. Bivariate LocusPlot illustrating the genome-wide association signals across chromosomal region 6:32682214-32897998 for Alzheimer’s disease (AD) and type 2 diabetes mellitus (T2DM). The left panel displays the −log10(p)-values from GWAS on T2DM (x-axis) contrasted with AD (y-axis), with each point corresponding to a single nucleotide polymorphism (SNP). The colour gradation for each SNP signifies the linkage disequilibrium (LD) with the lead SNP, as denoted by the colour scale. The right-hand plots emphasize the SNPs’ physical positions along chromosome 6 (x-axis) in relation to their −log10(p) (y-axis) for association with AD and T2DM. The lead SNP, identified as having a high posterior probability (PP) of being a shared causal variant, is distinctly accentuated in both plots. This highlights a genomic region of interest that may carry pleiotropic causal effects, potentially implicating this locus in the comorbidity of AD and T2DM.

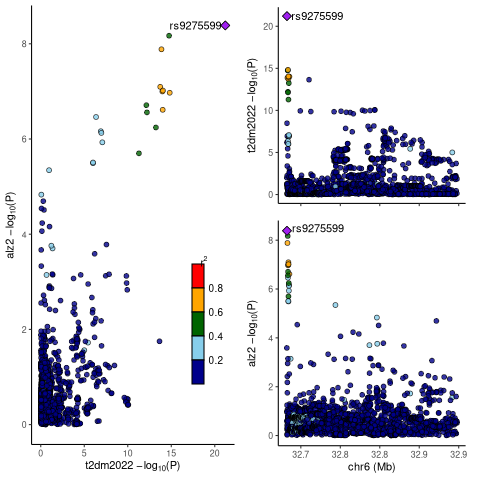
